# Refining Cell-free DNA Metagenomics for Sepsis in Acute Leukemia: Towards Standardized Computational Strategies for Host Depletion and Microbial Detection from Shallow-Depth, Low-Coverage Profiles

**DOI:** 10.1101/2025.10.17.25338171

**Authors:** Arpit Mathur, Karishma Anam, Sujata Lall, Elwin Paulose, Vishram Terse, Vaibhav Gawde, Prasanna Bhanshe, Swapnali Joshi, Shruti Chaudhary, Manisha More, Pratiksha Salunke, Prashant Tembhare, Sweta Rajpal, Gaurav Chatterjee, Papagudi Ganesan Subramanian, Sumeet Gujral, Sachin Punatar, Sumeet Mirgh, Akanksha Chichra, Gaurav Bain, Nishant Jindal, Lingaraj Nayak, Bhausaheb Bagal, Hasmukh Jain, Manju Sengar, Alok Shetty, Anant Gokarn, Vivek Bhatt, Navin Khattry, Nikhil Patkar

## Abstract

**Introduction:** Accurate alignment is critical for metagenomic profiling of plasma cell-free DNA (cfDNA), where microbial fragments are short, low in abundance, and often obscured by abundant human DNA. Yet, no unified guideline exists for selecting alignment algorithms in cfDNA studies.

**Results:** We benchmarked six alignment methods (BWA-MEM2, Minimap2, Bowtie2-sensitive, Bowtie2-very-sensitive, Bowtie2-very-sensitive-local, and Kraken2) using in silico simulated datasets and cfDNA from 102 acute leukemia samples with sepsis. Evaluation criteria included bacterial read retention after human read filtering, minimization of false positives in real cfDNA datasets, and detection sensitivity for antimicrobial resistance (AMR) genes. BWA-MEM2 emerged as the most effective for removing stringent human reads and bacterial classification, while bowtie2(very_sensitive_local mode) was optimal for AMR gene detection. Post-filtering, cfDNA profiles exhibited marked heterogeneity in bacterial and AMR gene signatures, reflecting expected clinical variability.

**Conclusions:** BWA-MEM2 provides optimal human read removal and bacterial taxa classification, minimizing false positives, though some true reads may be lost. For AMR gene detection, Bowtie2 (very_sensitive_local mode) demonstrated superior sensitivity in simulated datasets. This study presents the first systematic evaluation of read-based and k-mer–based aligners in plasma cfDNA, offering practical guidance for balancing human read removal with bacterial and AMR gene profiling to achieve robust and clinically meaningful results.

## Introduction

Metagenomics focuses on analyzing genomes beyond the human core genome, searching for signs of life, including bacteria, fungi, viruses, archaea, and other microorganisms, as well as their genetic functions, diversity, and interactions within a specific environment. Cell-free DNA (cfDNA) metagenomics is a specialized application where the microbial DNA is analyzed from fragmented DNA circulating freely in body fluids, typically plasma. Metagenomic analysis in cfDNA is particularly challenging due to various reasons, including low microbial biomass, ultra-sensitive short and fragmented DNA, High background of host (human) DNA, and the risk of contamination.

Blood cultures often miss infections, especially if samples are collected after antibiotic administration or when microbial load is low. Plasma cfDNA metagenomics can detect microbial DNA and AMR genes from viable or lysed pathogens when cultures are negative. Key resistance genes, such as *van A/B* and *mecA*, can be identified using this method^1^, providing critical insights for therapy and stewardship, particularly in critically ill patients^2,3^. cfDNA metagenomics offers a non-invasive, culture-independent method for uncovering sepsis and bacterial resistance.

While wet-lab methods have improved microbial cfDNA enrichment, computational challenges remain critical. Despite advances like host DNA depletion (Catalog Number:116545050, MP Biomedical) and size selection (Catalog Number: NG2001, Promega), no standard computational framework exists for contamination removal in cfDNA metagenomics. Low microbial biomass makes samples highly susceptible to reagents and lab impurities, which can skew results. While current tools, such as *Low biomass background correction* (LBBC)^4^, show promise, the authors did not benchmark their Bowtie2-based alignment strategy against other aligners. There is an urgent need for a robust benchmarking framework to comprehensively evaluate the capability of different aligners to help reliably distinguish true microbial signals from contamination across diverse cfDNA datasets.

Previous studies have highlighted the performance of Bowtie2 in human read removal from metagenomic samples. Forbes et al^5^ demonstrated that Bowtie2 in very-sensitive-local mode, combined with the CHM13-T2T reference, outperformed other aligners, achieving high sensitivity with minimal loss of specificity. Similarly, Constantinides et al^6^ reported superior performance using Bowtie2 in the standard sensitive mode with the CHM13-T2T reference. However, to date, there has been no comprehensive benchmarking of alignment algorithms specifically in the context of cell-free DNA (cfDNA) metagenomic samples. To the best of our knowledge, our study represents the first systematic evaluation of this kind in cfDNA.

Recent work by Abraham Gihawi et al^7^ highlighted significant data analysis errors that challenge previous findings in the cancer microbiome. They specifically reviewed a study by GD Poore et al^8^ and showed that many of the bacterial species reported were actually misidentified human DNA reads. This happened because of contamination in reference databases-some human DNA sequences were incorrectly labeled as bacterial, as draft bacterial genomes sometimes contain human DNA fragments that lead to misclassification.

An optimal alignment algorithm for filtering human-like reads and aligning cleaned data to bacterial or AMR gene databases remains uncertain, especially for plasma cfDNA. Our work aims to identify the best aligners for filtering human reads and accurately classifying bacterial and AMR gene sequences in cfDNA.

Custom reference databases enhance metagenomic classification by reducing false positives and improving taxonomic resolution, particularly in complex, host-associated samples. In contrast, standard databases often contain redundant or erroneous sequences that compromise precision. Incorporating host genomes into tailored databases further mitigates misclassification and increases specificity in cfDNA analyses^9,10^. Building on these principles, we performed alignment-based analysis of paired-end cfDNA profiles against known bacterial sequences using custom-built databases relevant to sepsis, thereby reducing false positives and improving true positive detection.

Published studies, as shown in the supplementary table, expose considerable variability in the combination of filtering and alignment algorithms, with the Bowtie2 filter being most frequently used, alongside numerous tools for alignment. For microbial alignment, Kraken2 is also widely used, while few studies employ Bowtie2, BWA, BLAST, or SNAP for both host and microbial assignments. In the current study, we address this gap by systematically benchmarking filtering and alignment algorithms to provide an evidence-based framework for cfDNA metagenomic analysis.

### Patient Recruitment & Sample Collection

Adult patients (≥18 years) with a confirmed diagnosis of Acute Leukemia (AL), classified according to the World Health Organization (WHO) 2017 criteria, and receiving induction chemotherapy or undergoing bone marrow transplantation (BMT), were prospectively enrolled following written informed consent. Inclusion criteria encompassed patients of either sex presenting with clinical features of febrile neutropenia or non-neutropenic sepsis during the course of therapy. Patients were excluded if they had hematologic malignancies other than AL, were receiving palliative care, or declined participation.

Peripheral blood specimens were collected at defined clinical timepoints: (i) at the onset of a febrile episode prior to initiation of empirical antimicrobial therapy, and/or (ii) at the time of any breakthrough fever during the same infectious episode, and/or (iii) 2^nd^ episode of febrile neutropenia/sepsis in the patient. A maximum of two sampling events per patient was permitted. Blood was drawn into Streck® Cell-Free DNA BCT® tubes (Streck, Inc., Omaha, NE, USA) to preserve circulating cell-free DNA integrity A simultaneous paired – peripheral blood and central line (whenever present) blood culture samples were collected in BacT/ALERT**®** culture bottles and BacT/ALERT**®** 3D automated blood culture microbial detection system (bioMérieux, Marcy l’Étoile, France) was used for growth and detection of micro-organisms. Blood biochemistry and blood counts were also evaluated. Sepsis-related markers, e.g., C-reactive protein and procalcitonin, were ordered as per the treating physician’s discretion. Imaging, e.g., X-rays or Computed Tomogram of the chest of patients, was done as per clinical condition whenever indicated.

All samples were processed under aseptic conditions, and negative template controls (NTCs) were incorporated throughout the workflow to monitor for potential exogenous DNA contamination during downstream library preparation and sequencing procedures.

### Circulating Cell-Free (ccf) DNA Extraction, Library Preparation & Sequencing

Peripheral blood samples were collected in Streck® Cell-Free DNA BCT® tubes Streck® Cell-Free DNA BCT® tubes (Streck, Inc., Omaha, NE, USA, Catalogue no. 230471), which stabilize nucleated blood cells and inhibit cellular lysis^11^, thereby preserving the integrity of circulating cfDNA during sample storage, transport, and batching. Following collection, samples were centrifuged at 2,500 rpm for 10 minutes at room temperature to separate plasma from cellular components. cfDNA was extracted from plasma using the Maxwell® RSC cfDNA Plasma Kit Maxwell® RSC cfDNA Plasma Kit (Promega, Madison, WI, USA, Catalogue no. AS1480) on the automated Maxwell® RSC Instrument, following the manufacturer’s protocol (AS1480). Quantification of extracted cfDNA was performed using the Qubit™ 1X dsDNA High Sensitivity Assay Kit (Thermo Fisher Scientific, Waltham, USA, Catalog no. Q33230). Purified cfDNA samples were stored at 4°C until further processing.

Libraries were prepared from 0.5–1 ng of cfDNA using the xGen™ Single-Stranded DNA & Low-Input DNA Library Preparation Kit (Integrated DNA Technologies, USA, Catalogue no 10009817), according to the manufacturer’s protocol, with a minor modification: the final bead cleanup elution was performed in 30 µL of low-EDTA TE buffer to enhance cfDNA stability and downstream compatibility. The library preparation workflow included enzymatic fragmentation, end repair, adaptor ligation, and single-strand library conversion optimized for low-input cfDNA. Library quality and concentration were assessed with the help of Qubit™ 1X dsDNA High Sensitivity Assay Kit (Thermo Fisher Scientific, Waltham, USA, Catalogue no. Q33230) and TapeStation (Bioanalyzer 4200, Agilent Technologies, Santa Clara, CA, USA) before sequencing.

Prepared libraries were sequenced on the Illumina NextSeq 1000 and NovaSeq 6000 platforms, generating paired-end reads of 300 bp. Each sample yielded an average of ∼42.48 million reads (150 x 2), with an average coverage of 3.583x, enabling high-resolution, untargeted metagenomic analysis of the circulating cfDNA. Five negative template controls (NTCs) were included in the sequencing runs to monitor contamination and assess potential batch effects. A total of 102 samples from 74 patients were sequenced. Among these, 28 patients experienced two episodes of fever, contributing a total of 56 samples, while the remaining 46 patients had a single episode of sepsis, yielding 46 samples. Additionally, one biologically negative control (BNC) sample was sequenced to establish baseline signals from AL patients without sepsis.

### Bioinformatics Methodology

To systematically evaluate and identify the most suitable aligner for host read removal in plasma cfDNA from sepsis patients, we developed a three-tier benchmarking strategy (Figure 1). This framework assesses each aligner across three critical domains: retention (ability to preserve true microbial reads), stringency (efficiency in removing host-derived reads), and specificity (minimizing residual human sequences after filtering). Together, these complementary evaluations provide a comprehensive basis for selecting the best-performing aligner.

**Figure No. 1:**
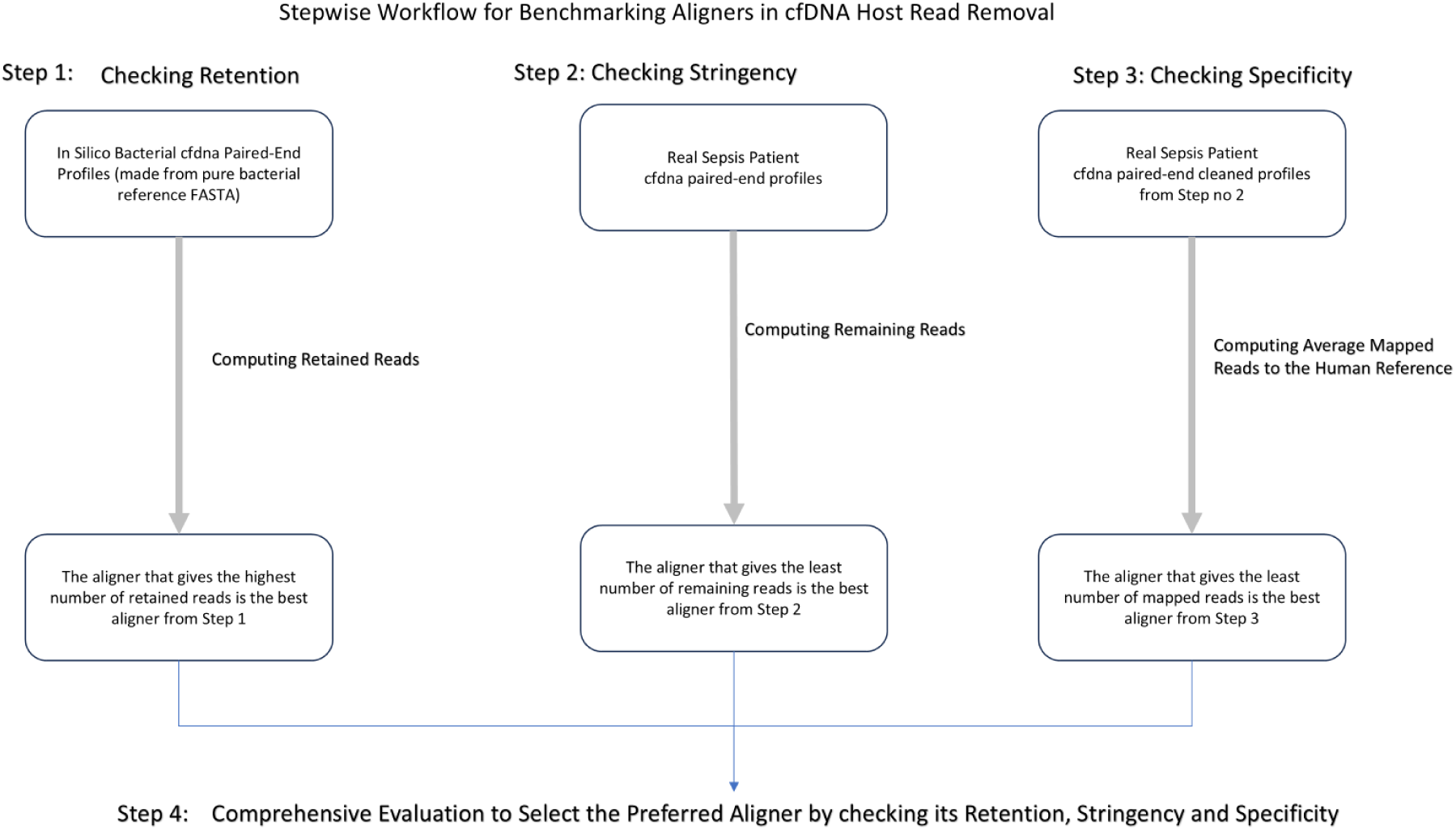
Three-Step Evaluation Framework for Benchmarking Aligners in cfDNA Host Decontamination

### (1) Pre-Processing of data

We trimmed Illumina primers/adapters from raw paired-end reads (Q20 cutoff), generated FASTQC reports pre- and post-filtering, and removed fragments <45 bp using *fastp* to exclude adapter dimers, low-quality reads, and ultra-short cfDNA fragments unlikely to represent true biological signal^12^ For the entire bioinformatics methodology, high-performance minimap2 was taken from mm2-fast^13^, bwa-mem, the latest version^14^ was taken, bowtie2 (all three modes) was taken from Ben Langmead et al^15^, while kraken2 was used from Derric E Wood et al^16^. For all aligners, their respective indices were created using the reference fasta of GCF_009914755.1^17^, which is the standard reference genome for CHM13T2T. To create the Kraken2 index, we developed a custom GitHub Tool located at https://github.com/arpit20328/FastaKrakenizer. This tool converts any custom FASTA file into a Kraken2 index without requiring hierarchy files (names.dmp and nodes.dmp) or an accession list. FastaKrakenizer makes dummy names, dmp, and nodes.dmp files treating each fasta header as a species alone, and thereby reporting every matched header in its report as a species matched. The Karken2 index of GCF_009914755.1 that was used in our study is available on the Zenodo records at https://zenodo.org/records/16459107.

An essential requirement of any cfDNA filtering algorithm is that it should efficiently remove human reads without discarding the target microbial reads. To evaluate this, we assessed which alignment algorithm best balances human read removal with bacterial read retention. To test this hypothesis, we created in-silico controls from bacterial reference fasta sequences, which were then tested using various alignment algorithms to check on their bacterial read retention capability.

### (2) Step 1: Checking Retention: Simulation-Based Assessment of Aligner Strategies for cfDNA Filtering

For the creation of in-silico paired-end profiles mimicking the cfDNA nature of our patient cohort, we used bacterial reference genomes from the NCBI database from its official Github pipeline named datasets^18^. We downloaded 23 reference genomes of bacteria, which are known to be the most common cause of bloodstream sepsis (supplementary table 1). To create an in-silico profile that mimics cfDNA data of our patient cohort, we used *InSilicoSeq*^19^, which generates customized paired-end fastq data.

For a given species (e.g., *X*), its reference genome was downloaded from NCBI. cfDNA reads were then simulated using *InSilicoSeq*^19^ with parameters chosen to match the fragment length distribution, read length, and coverage observed in patient cfDNA samples (169 bp mean fragment length, 26 bp standard deviation, 71,319,000 paired-end reads, NovaSeq model). Generated reads were subsequently processed with fastp to remove fragments shorter than 45 bp, as described above.

We evaluated six aligners for human read removal, as the optimal choice for each bacterial species was not known beforehand. For each species, we calculated the median number of reads retained across aligners, using the median instead of the mean to reduce the influence of outliers. The aligner whose retained read count was closest to this median was considered the best performer, as it most accurately preserved the expected proportion of non-human (bacterial) reads. Reads in the original dataset and reads remaining in each aligner case for each species are provided in Supplementary Table No. 1.

### (3) Step 2: Checking Stringency: Assessment of Residual Reads After Filtering

To come to a conclusion, as to which algorithm is best for human read removal, we took random fifteen cfDNA fastq paired-end FASTQ profiles from our patient cohort and cleaned the data by removing human reads with the use of human reference fasta GCF_009914755.1 using all six aligners as previously performed in the Supplementary Table No. 1 exercise.

Table No. 1 shows the remaining read count in million (M) remaining after filtering human reads for every patient when human reads are filtered via six different aligners.

**Table No. 1.**
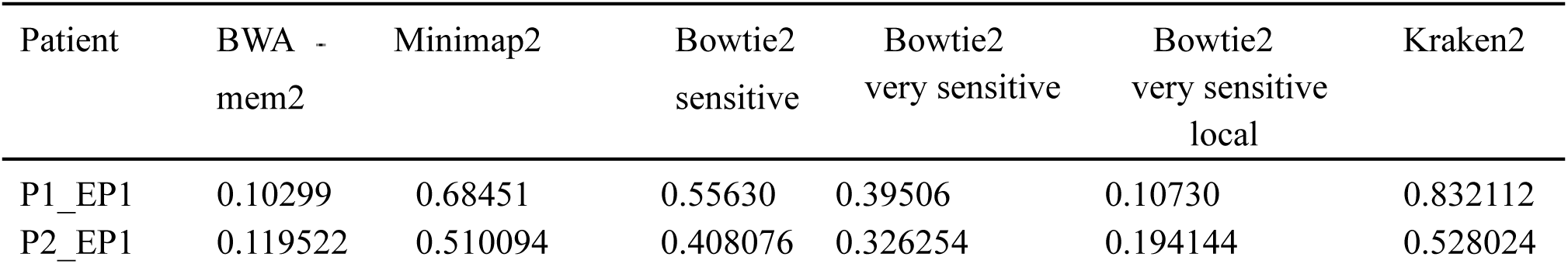
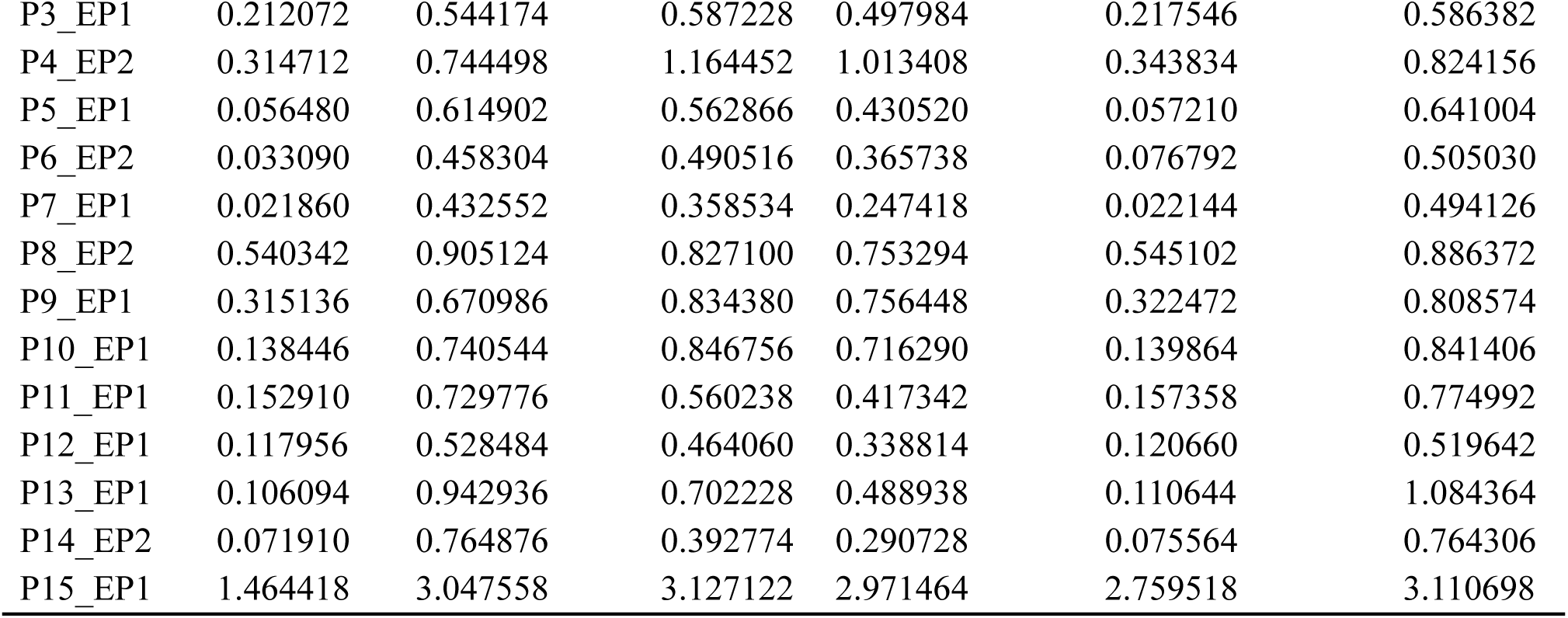
Comparison of Remaining Reads (M) Across Six Aligners for Human Read Removal in Patient Samples.

| Patient | BWA -<br>mem2 | Minimap2 | Bowtie2<br>sensitive | Bowtie2<br>very sensitive | Bowtie2<br>very sensitive<br>local | Kraken2 |
| --- | --- | --- | --- | --- | --- | --- |
| P1_EP1 | 0.10299 | 0.68451 | 0.55630 | 0.39506 | 0.10730 | 0.832112 |
| P2_EP1 | 0.119522 | 0.510094 | 0.408076 | 0.326254 | 0.194144 | 0.528024 |
| P3_EP1 | 0.212072 | 0.544174 | 0.587228 | 0.497984 | 0.217546 | 0.586382 |
| P4_EP2 | 0.314712 | 0.744498 | 1.164452 | 1.013408 | 0.343834 | 0.824156 |
| P5_EP1 | 0.056480 | 0.614902 | 0.562866 | 0.430520 | 0.057210 | 0.641004 |
| P6_EP2 | 0.033090 | 0.458304 | 0.490516 | 0.365738 | 0.076792 | 0.505030 |
| P7_EP1 | 0.021860 | 0.432552 | 0.358534 | 0.247418 | 0.022144 | 0.494126 |
| P8_EP2 | 0.540342 | 0.905124 | 0.827100 | 0.753294 | 0.545102 | 0.886372 |
| P9_EP1 | 0.315136 | 0.670986 | 0.834380 | 0.756448 | 0.322472 | 0.808574 |
| P10_EP1 | 0.138446 | 0.740544 | 0.846756 | 0.716290 | 0.139864 | 0.841406 |
| P11_EP1 | 0.152910 | 0.729776 | 0.560238 | 0.417342 | 0.157358 | 0.774992 |
| P12_EP1 | 0.117956 | 0.528484 | 0.464060 | 0.338814 | 0.120660 | 0.519642 |
| P13_EP1 | 0.106094 | 0.942936 | 0.702228 | 0.488938 | 0.110644 | 1.084364 |
| P14_EP2 | 0.071910 | 0.764876 | 0.392774 | 0.290728 | 0.075564 | 0.764306 |
| P15_EP1 | 1.464418 | 3.047558 | 3.127122 | 2.971464 | 2.759518 | 3.110698 |

### (4) Step 3: Checking Specificity: Assessment of Residual Human Reads After Filtering

To evaluate the efficiency of each aligner in removing human reads, we analyzed the residual DNA content in the filtered cfDNA datasets. For each patient, we started with six filtered read sets—each obtained after applying one of the six filtering aligners. We then re-aligned each of these six filtered datasets against the human reference genome (GCF_009914755.1) using all six aligners. This resulted in 36 total alignments per patient (6 filtered datasets × 6 aligners).

We then calculated the median number of reads mapped to the human reference across these six aligners for each filtered dataset. This approach provides a robust, unbiased estimate of the amount of residual human contamination remaining after filtering, accounting for differences in aligner stringency and sensitivity. Aligners that yield fewer mapped human reads after filtering are considered more effective in removing human sequences. The aggregated results of this comprehensive cross-alignment analysis are presented in Table No. 2.

**Table No. 2.** Residual Human Read Counts After Filtering: Cross-Alignment Comparison Across Six Aligners.

| Sample | Bowtie2<br>Sensitive | Bowtie2<br>Very-Sensitive | Bowtie2<br>Very-Sensitive-Local | BWA-MEM2 | Kraken2 | Minimap2 |
| --- | --- | --- | --- | --- | --- | --- |
| P1_EP1 | 0.364223 | 0.235747 | 0.020845 | 0.019758 | 0.583009 | 0.496440 |
| P2_EP1 | 0.193787 | 0.126182 | 0.029145 | 0.010151 | 0.266869 | 0.289607 |
| P3_EP1 | 0.259996 | 0.191892 | 0.020528 | 0.019344 | 0.304999 | 0.254829 |
| P4_EP2 | 0.615888 | 0.494044 | 0.063395 | 0.057361 | 0.373732 | 0.346276 |
| P5_EP1 | 0.389582 | 0.277270 | 0.012075 | 0.011994 | 0.465857 | 0.466892 |
| P6_EP2 | 0.320391 | 0.217083 | 0.014526 | 0.003016 | 0.339390 | 0.322490 |
| P7_EP1 | 0.272292 | 0.177309 | 0.005140 | 0.005129 | 0.381817 | 0.343355 |
| P8_EP2 | 0.313927 | 0.251189 | 0.105015 | 0.104192 | 0.328513 | 0.398463 |
| P9_EP1 | 0.408678 | 0.343819 | 0.059357 | 0.058929 | 0.391668 | 0.328101 |
| P10_EP1 | 0.525640 | 0.416422 | 0.027139 | 0.028054 | 0.549196 | 0.504185 |
| P11_EP1 | 0.329417 | 0.225408 | 0.031029 | 0.029894 | 0.496221 | 0.503336 |
| P12_EP1 | 0.288903 | 0.183720 | 0.021350 | 0.020710 | 0.319748 | 0.357484 |
| P13_EP1 | 0.477428 | 0.303992 | 0.021761 | 0.020652 | 0.785191 | 0.708058 |
| P14_EP2 | 0.257563 | 0.169983 | 0.015431 | 0.014847 | 0.561074 | 0.579673 |
| P15_EP1 | 0.723535 | 0.599398 | 0.447419 | 0.110033 | 0.276176 | 0.684103 |

We additionally assessed stringency and specificity using three public cell-free DNA datasets (PRJNA704313^20^, PRJNA742001^21^, and PRJNA782906^22^), comprising a total of 202 sepsis samples.

### (5) Curated Bacterial Database for Sepsis

The bacterial reference database used in this study was derived from the pathogenic database developed by Nikolay Oskolkov et al^23^, representing a curated subset of NCBI non-redundant NT/NR records as of May 2020. Only species listed in the “Sepsis Database” (except Candida and Lactobacillus) in the Supplementary Sheet were retained, as these are the common causative organisms of sepsis in patients with febrile neutropenia. To improve taxonomic resolution, we additionally incorporated 16S rRNA sequences from the EMU pipeline default database^24^, again selecting only species corresponding to “Sepsis Database”. The resulting combined resource, referred to hereafter as the “Combined Filtered D1 Database”, is publicly available at Zenodo records at https://zenodo.org/records/16721528.

### (6) Targeted vs. Complete Human DNA Removal Without Targeted Masking

We analyzed a “Combined Filtered D1 Database” FASTA bacterial sequences against a human reference genome of 24 sequences (GCF_009914755.1_T2T-CHM13v2.0). Average Nucleotide Identity (ANI) analysis revealed that only 20 query sequences had ∼70% similarity to human DNA, while the rest showed low similarity. We therefore applied a two-step human DNA removal strategy. First, all human sequences were removed without targeted masking of bacterial DNA, ensuring complete depletion of human reads. Second, targeted filtering retained bacterial sequences of interest, minimizing loss of genuine microbial DNA. Avoiding targeted masking first prevents residual human-like reads from remaining, ensuring maximal bacterial sequence preservation and reliable downstream analyses, including per-sequence ANI estimation and microbial profiling.

### (7) Selection of Alignment Algorithm for Post-Filtering Taxa Identification

To assess alignment algorithm accuracy, we used 23 previously generated *in silico* cfDNA profiles of bacterial species (“Sepsis Database” in the Supplementary Sheet). Patient-derived filtered cfDNA data (n = 15) had an average insert size of 150 bp and an average of 0.110 million paired-end reads. Accordingly, for each of the 23 reference genomes, we simulated cfDNA-like paired-end reads using InSilicoSeq^19^ (NovaSeq model; 220,704 reads; mean fragment length: 150 bp; standard deviation: 50 bp) to match patient cfDNA characteristics. Simulated reads were quality filtered with fastp, retaining only fragments ≥45 bp.

These profiles were subsampled to 0.2622 million reads, which corresponds to the average number of reads remaining after filtering with BWA-MEM2 for 15 patient cfDNA profiles (Table 1). To evaluate aligner performance, synthetic cfDNA reads were simulated from known bacterial reference genomes (Supplementary Table 1) and aligned to our custom “Combined Filtered D1 Database”, with high-confidence alignments retained at MAPQ ≥30 and a confidence score of 0 and above for kraken2 alignment.

Applying stringent mapping quality filters highlights the trade-off between precision and recall in metagenomic analyses. For instance, benchmarking with the *EukDetect*^25^ marker gene database, the authors demonstrated that a MAPQ ≥ 30 cutoff increased precision from 95.1% to 99.7% by eliminating spurious alignments, but reduced recall to 91.7% by discarding true reads mapping to conserved regions. Thus, higher thresholds improve specificity at the expense of sensitivity, underscoring the need for context-dependent parameter selection in cfDNA studies. In our study, we have taken MAPQ greater than or equal to 30 for all downstream analysis.

In this study, true hits were defined as reads aligning correctly to the complete genome or full-length 16S rRNA gene sequences of the expected bacterial species. False hits included reads mapping to plasmids, partial gene sequences, insertion elements, or incorrect species/genomes. Based on these definitions, we quantified alignment performance using standard metrics: true positives (TP), false positives (FP), and false negatives (FN).

From these metrics, we calculated precision, recall, the F1 score, and Mapping efficiency, representing the proportion of all reads mapped (true or false hits), which was also computed according to the formulas in the supplementary tab formulae. This framework provides a rigorous benchmark for evaluating aligner accuracy in high-resolution metagenomic analyses, with direct relevance to clinical diagnostics and pathogen surveillance.

Figure No.2: Diagram illustrating the distribution of reads from total input through alignment outcomes. The plot highlights how aligned reads are classified into true hits (e.g., complete genomes, full-length 16S) and false hits (e.g., plasmids, partial genes, insertion elements, or incorrect species), showcasing an aligner’s performance fidelity. Complete aligner information for each of the 23 reference species is shown in the Supplementary Sheet.

**Figure 2:**
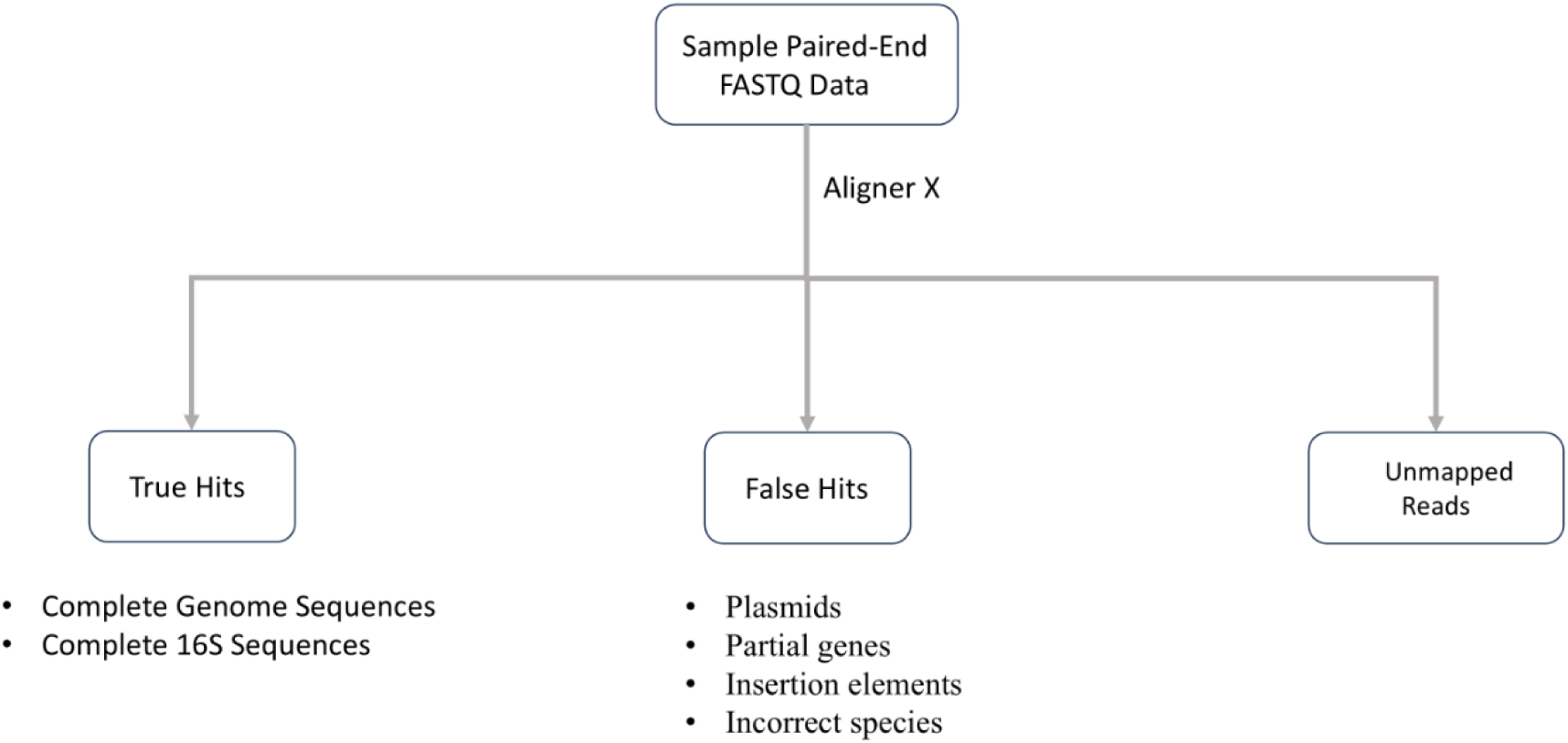
Read Flow from Input to Alignment Accuracy: Diagram of True vs. False Hits for Aligner “X"

This analysis also enabled us to assess the coverage characteristics of true alignments. For each true positive (TP), we measured the breadth and depth of coverage across the originating reference. By examining these metrics, we identified the minimum coverage thresholds typically associated with accurate detection. This provides a valuable baseline to interpret real patient cfDNA data, allowing us to estimate the minimum breadth and depth at which a species-level call can be considered reliable.

To identify the optimal aligner for each bacterial species, we used the F1 score as the primary ranking metric. The F1 score, defined as the harmonic mean of Precision and Recall, provides a balanced measure of an aligner’s ability to correctly identify accurate microbial reads while minimizing both false positives and false negatives. For each species, all aligners were evaluated based on their computed F1 scores, and the aligner achieving the highest F1 score was designated as the best-performing tool. In instances where two or more aligners had identical F1 scores, Precision and Mapping Efficiency were applied as secondary criteria to resolve ties, favoring aligners that made fewer incorrect assignments and mapped the largest proportion of reads. This ranking framework enabled objective, species-specific benchmarking of aligners, emphasizing accuracy in high-resolution metagenomic analyses. The results are summarized in the Supplementary sheet, which lists the best-performing aligner for each species.

### (8) Criteria for Reporting Bacterial Pathogens in Plasma cfDNA Metagenomic Sequencing

We reviewed literature to understand how different studies defined and reported bacterial pathogens from broad metagenomic next-generation sequencing (mNGS) results of plasma cfDNA. Each group applied a combination of quantitative thresholds, comparison with negative controls, and clinical adjudication to distinguish true pathogens from background signals or contaminants. A summary of the reporting strategies is provided in the following Table No. 3.

**Table No. 3:** Reporting Strategies for Bacterial Pathogens in Plasma cfDNA mNGS.

| Study | Read/Abundance Threshold |
| --- | --- |
| Hui Zhang et al,<sup>26</sup> (2024) | · The genome coverage of uniquely mapped reads placed this microorganism among the top 10 microbes of the same type, and it was absent in the negative control (NTC).<br>· The ratio of RPM in the sample to RPM in the NTC exceeded 10 (with RPM in the NTC not equal to zero). |
| Qiuyu Jing et al<sup>27</sup> (2022) | · Confirmed by a minimum of 10 supporting reads.<br>· The read count in the sample was at least twice that observed in the NTC.<br>· Displayed a relative abundance of at least 1%. |
| Chunhui Xu et al<sup>28</sup> (2023) | · For bacterial and fungal taxa, the genus-level relative abundance was required to exceed 30%.<br>· For bacteria, a species was classified as positive if the stringently mapped read number (SMRN) was at least 3.<br>· Positive microbial detections had to show a higher SMRN than that observed in the negative controls. |
| Thomas Schmoch et al<sup>29</sup> (2021) | · The abundance of different species in the microbiological samples will be evaluated using the Sepsis Indicating Quantifier (SIQ) score |
| Chaoqin Zhang et al<sup>30</sup> (2022) | · mNGS detected a species-level microbe with a coverage rate that was at least ten times higher than that of any other identified microbes. |

Since there are no universally accepted guidelines for reporting pathogenic organisms in plasma cfDNA mNGS, we adopted consensus criteria from prior studies. Pathogen detection from mNGS data was performed using a multi-step filtering approach designed to minimize false positives. Species were first required to have at least 10 uniquely mapped reads. To account for background signals, any species also detected in negative controls (NTCs) was retained in a sample only if its read count was at least two-fold higher, its absolute abundance was at least two-fold higher, and its genome coverage fraction was at least ten-fold higher than in the corresponding NTC. NTC-derived signals were considered for contamination only if the species was present in at least two NTCs, while species detected in a biologically negative control (BNC) were directly included as controls. Following NTC filtering, species with a relative abundance greater than 1% were considered for further analysis.

### Benchmarking the Best Alignment Algorithm for AMR Gene Detection from Cleaned cfDNA Data

To identify the most accurate aligner for cleaned cfDNA data (depleted of human reads), we analyzed in-silico cfDNA profiles constructed from 35 reference bacterial genomes containing known AMR genotypes from NIH Isolate browser^31^ (listed in Supplementary AMR gene In-silico database). These profiles were aligned using the same six aligners, and the ratio of true to false hits was calculated. True hits were defined as read mapping to the known AMR genes of the specific species, while false hits were read mapping to AMR genes (specific to species) not present in the corresponding reference genotype. The aligner yielding the highest true-to-false hit ratio was selected as the optimal choice. The CARD database^32^ was used for this analysis.

For reporting AMR genes of high-confidence true calls, reads having more than 95% identity were considered as reported in Mostafa M. H. Ellabaan et al^33^, with more than five reads and having a fraction of genome coverage more than 10% as suggested in Dan Lu et al^34^ were considered as true hits. If AMR genes were detected in both NTCs and patient samples, lab contamination was controlled by requiring that the gene in the sample have >2× the read count, >2× the absolute abundance, and >10× the fraction of genome covered compared with the NTCs. NTC-derived signals were considered only if the gene was present in at least two NTCs, whereas species detected in the biologically negative control (BNC) were directly included as controls.

## Results

### (1) Retention

Across 23 in-silico bacterial cfDNA profiles (Supplementary Table 1), Bowtie2 Sensitive consistently retained microbial reads closest to the original total, with Very Sensitive performing optimally in most species. Very Sensitive Local showed moderate but never superior performance. BWA-MEM2 under-retained reads due to stringent filtering, while Minimap2 and Kraken2 were variable and less consistent. Overall, Bowtie2’s sensitive modes provided the best balance between human read removal and microbial read preservation, a trend maintained under stricter MAPQ thresholds.

### (2) Stringency

Post-filtering read counts from fifteen cfDNA samples (Table 1) showed BWA-MEM2 consistently left the fewest human reads (0.02M–1.46M), indicating superior host depletion. Bowtie2_VerySensitiveLocal and Bowtie2_VerySensitive ranked next, while Minimap2 and Kraken2 were intermediate. Bowtie2_Sensitive often retained the most human reads, reflecting weaker filtering. Similar trends held under stricter MAPQ thresholds.

Stringency was tested on three cfdna public databases (sepsis cohort) as listed in the supplementary sheet. A total of 202 sepsis samples, stringency was checked with similar aligners, and bwa-mem2 was the most optimal stringent aligner in 201 cases.

### (3) Specificity

Table 2 shows that BWA-MEM2 and Bowtie2 (very-sensitive-local) yielded the lowest residual human reads (∼0.03–0.11M; >99.5% removal), while Bowtie2 (sensitive/very-sensitive) retained moderate levels, and Minimap2/Kraken2 showed higher residuals (>0.30M). Across 15 patients, BWA-MEM2 was most stringent in 14 cases.

Specificity tested on a total of 202 samples from three public sepsis cfdna studies revealed BWA-MEM2 as the most specific aligner in 188 samples. (results in supplementary sheet)

### (4) Integrated Evaluation of Aligner Stringency and Microbial Retention

Human read removal in cfDNA requires a careful balance between stringency and specificity. Less stringent aligners may retain residual human reads, leading to false positives, whereas highly stringent aligners risk discarding genuine microbial sequences. As shown in Figure 3, BWA-MEM2 achieved the optimal balance, effectively removing nearly all human sequences while preserving true microbial and AMR gene signals.

**Figure No. 3:**
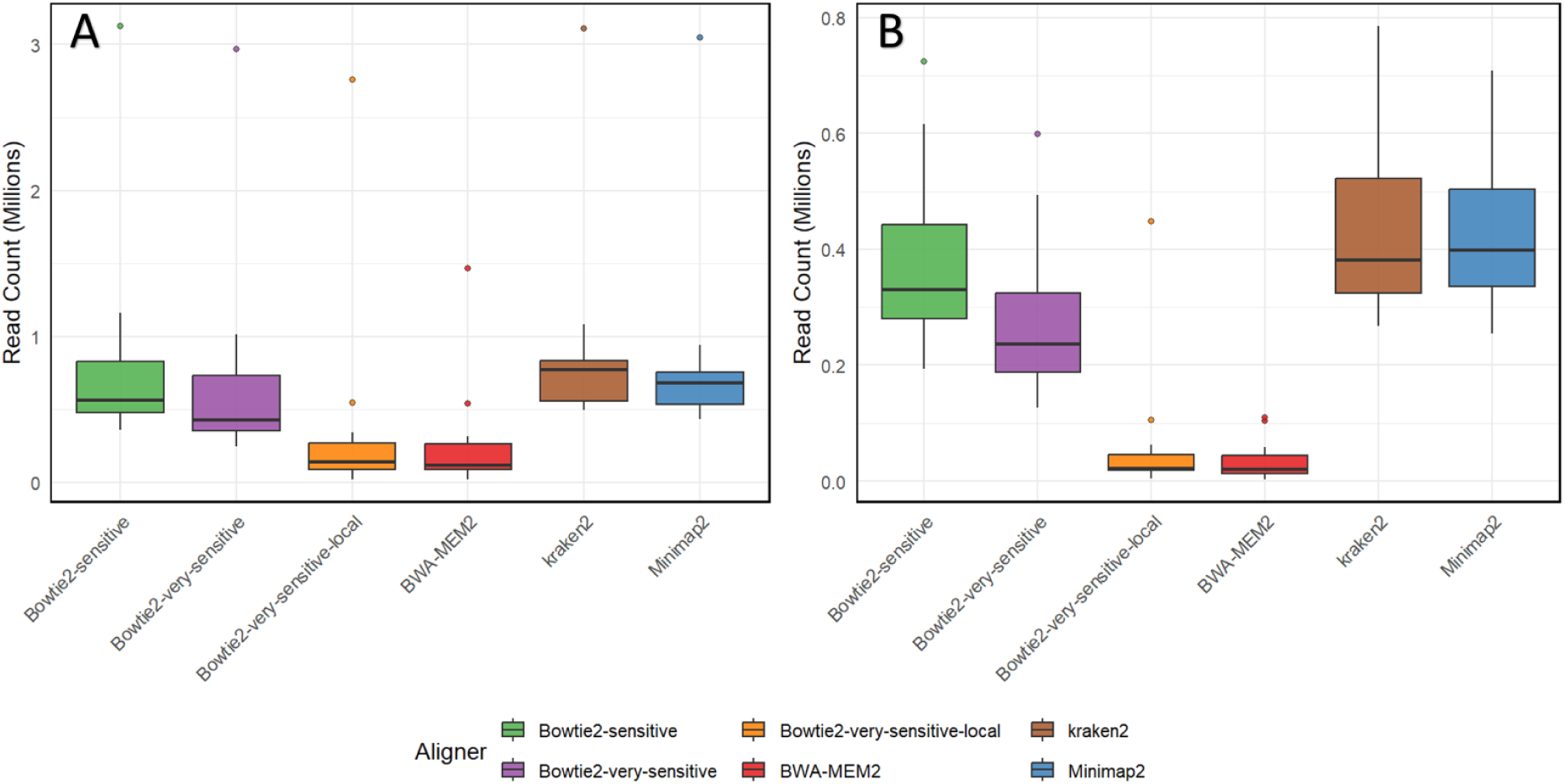
Boxplot Analysis of Post-Filtering Retained Reads (Fig. A) and Residual Human Reads (Fig. B) Across Six Aligners

When examining the ratio of human reads (from Table 2) to remaining reads (from Table 1), BWA-MEM2 had the lowest ratio in 9 out of 15 patients, while Bowtie2 (very-sensitive local) achieved the lowest ratio in the remaining 6 cases. The superior performance of BWA-MEM2 was further confirmed using in-silico cfDNA profiles, where it successfully extracted all human reads, demonstrating its stringent yet reliable filtering capability.

### (5) Viral Contamination

Contamination from viral reads was low (∼4.65%), and given the breadth of NCBI RefSeq, the true plasma-relevant viral content is likely even lower. More stringent MAPQ and Kraken2 thresholds further reduced viral reads, confirming that the data were sufficiently clean for accurate bacterial classification and AMR gene detection. A detailed viral removal strategy would require additional benchmarking and is beyond the scope of this study (see Supplementary Viral Alignment Statistics).

### (6) Aligner for Filtering Human Reads

Based on the above results, we designed our filtering strategy for removing human reads from cfDNA plasma profiles using the BWA-MEM2 aligner.

### (7) Aligner for Post Filtering Bacterial Taxa Identification

In our D1 dataset, complete genome FASTA sequences for the following five species—Providencia rettgeri, Elizabethkingia meningoseptica, Candida albicans, Sphingomonas paucimobilis, and Bacillus subtilis—were excluded from downstream analysis. After this filtering step, 18 species remained for robust comparative evaluation of aligner accuracy and efficiency.

Across the 18 bacterial species evaluated, BWA-MEM2 demonstrated the strongest overall performance, achieving an average F1 score of 0.27, with an average precision of 0.49 and a mapping efficiency of 35.24% across these 18 species. This indicates that BWA-MEM2 consistently identified accurate reads while minimizing incorrect assignments for the majority of species. In contrast, Minimap2 showed lower performance, with a mean F1 score of 0.15, followed by kraken2 and bowtie2 (very-sensitive-mode).

These results indicate that BWA-MEM2 is the most robust and accurate aligner overall, providing a favorable balance between precision and recall across a broad range of bacterial species. In contrast, Minimap2 exhibits limited sensitivity and mapping efficiency, particularly for low-abundance or complex genomes, highlighting the critical importance of aligner selection in high-resolution metagenomic analyses (Table No. 5).

**Table No.5:** Overall Performance Metrics of Aligner Tools After Filtering Low True-Positive Species.

| Rank | Aligner | Precision | Recall | F1_Score | Mapping Efficiency (%) |
| --- | --- | --- | --- | --- | --- |
| 1 | bwa_mem2 | 0.498 | 0.2267 | 0.2726 | 35.2428 |
| 2 | minimap2 | 0.4485 | 0.096 | 0.1578 | 17.1632 |
| 3 | kraken2 | 0.1912 | 0.0044 | 0.0082 | 2.026 |
| 4 | bowtie2<br>(very_sensitive ) | 0.003 | 0 | 0 | 0.362 |

Based on the analyses presented in Figure 3 and Table No. 5, BWA-MEM2 emerged as the most effective aligner for removing human reads while preserving targeted microbial sequences. It is also the preferred tool for taxonomic classification of filtered cell-free DNA profiles, as illustrated in Figure 4.

**Figure No. 4:**
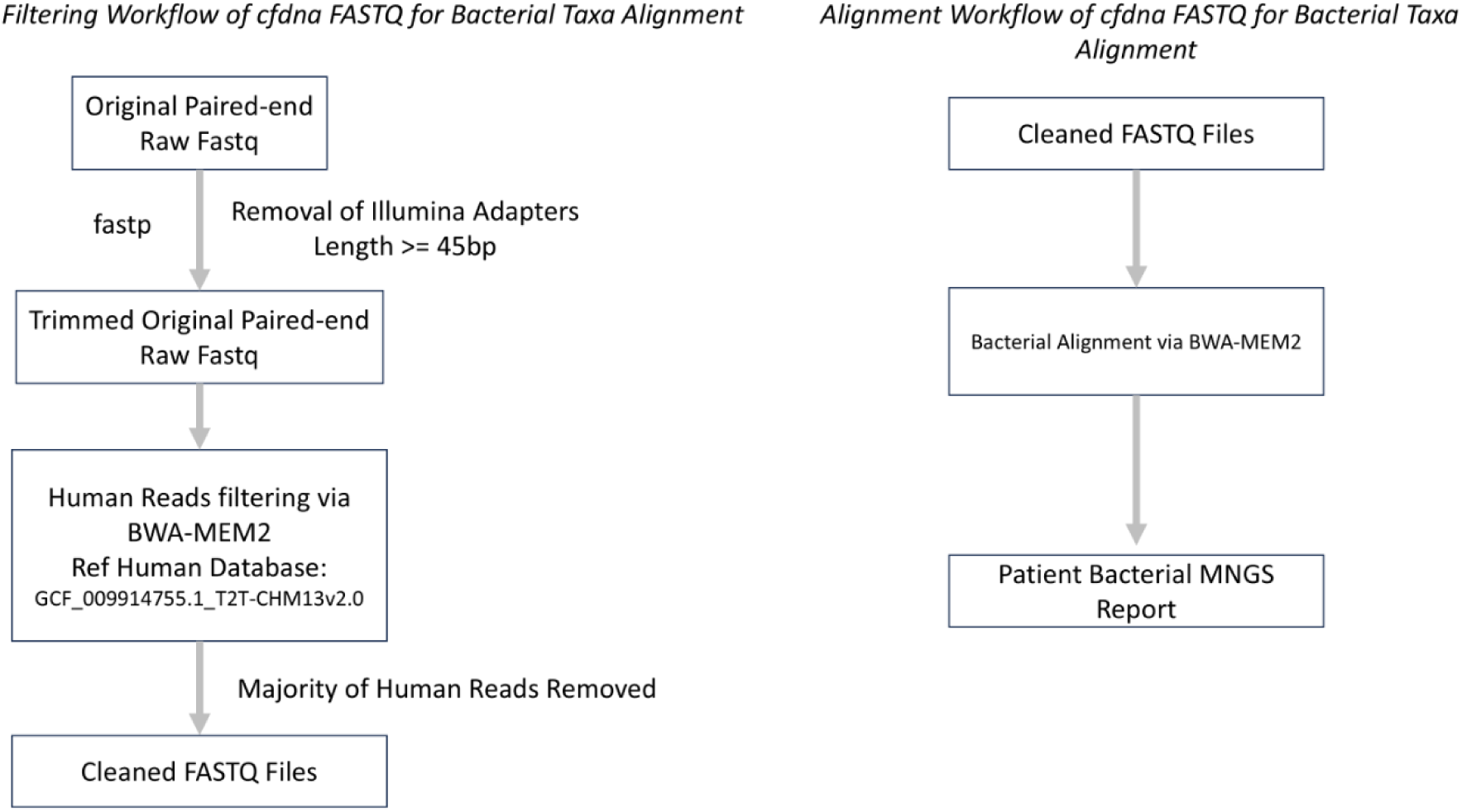
Recommended Aligners for cfDNA Analysis: Host Depletion and Taxonomic Classification

### (8) Result of Bacterial Taxa Alignment post filtering

Before human read decontamination, 68 samples out of 102 had at least one species present, while after removal of human reads, 85 samples had bacterial species presence. Before filtering the top 5 frequent species, *Sphingomonas paucimobilis* had a dominant presence (24 samples), *Staphylococcus aureus isolates* (15 samples), *Elizabethkingia meningoseptica* (13 samples), *Escherichia coli strain BCRC* (9 samples), and *Enterobacter hormaechei subsp. Oharae strain* (8 samples) was completely absent post-human read removal. Alternatively, the most frequent species after filtering, like Bacillus cereus G9842 (in 38 samples), Stenotrophomonas maltophilia K279a (31 samples), etc, were absent in the pre-filtering scenario across 102 samples.

Further, there was no single sample that had a similar microbial spectrum pre- and post-human read step, demonstrating the importance of the bioinformatic human read step and its impact on microbial profiling of samples, which can lead to false clinical and biological interpretations. The Pheatmap showing changes in the presence/absence of bacterial species-pre and post-human-removal step is shown below in Figure 5. The individual and comparative frequency table of bacterial species in both pre- and post-human read filtering steps, per sample, is in the supplementary.

**Figure 5:**
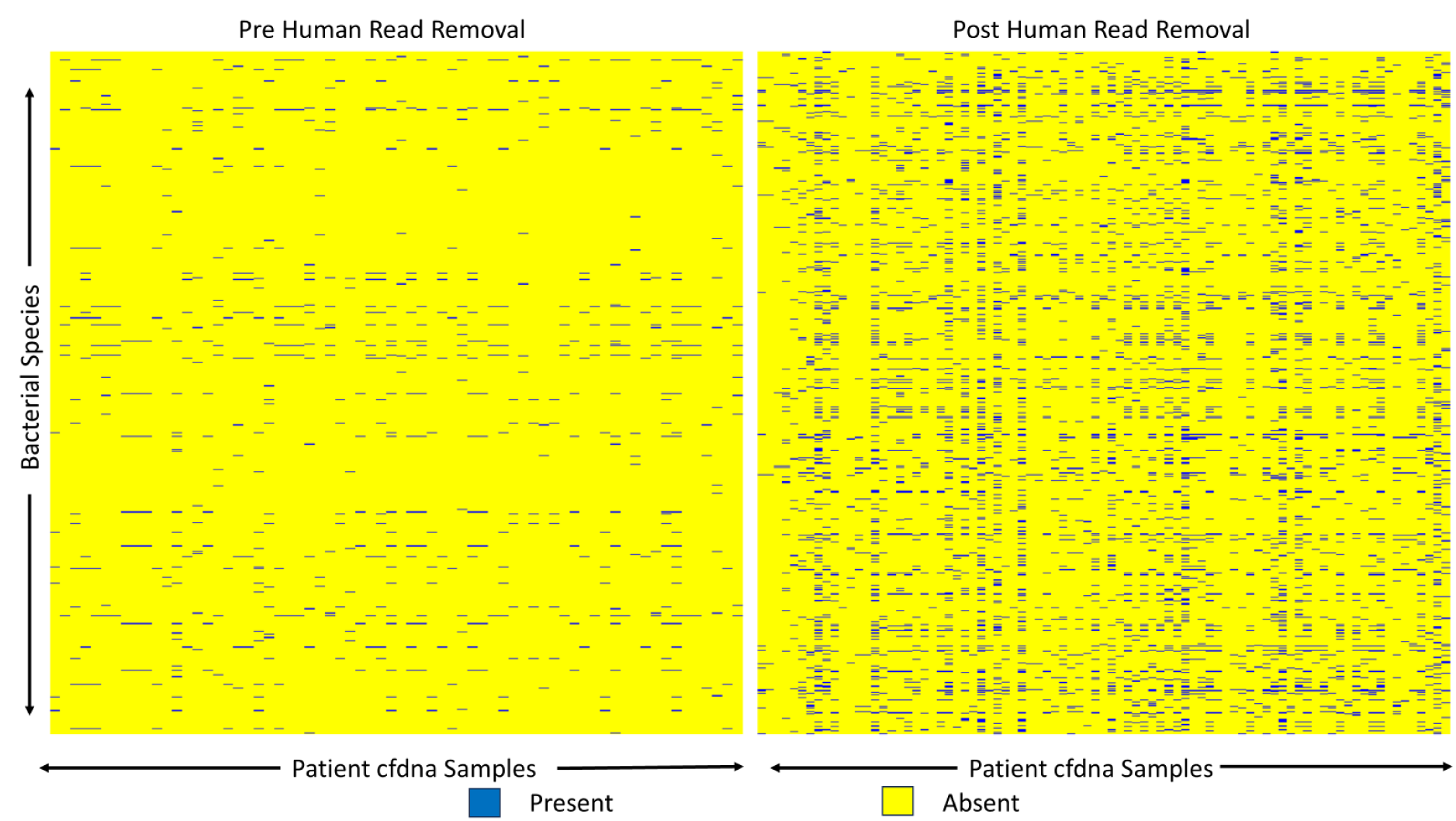
Pheatmap of Bacterial Species presence before and after human read filtering over all 102 cfdna samples.

### (9) Aligner for Post Filtering AMR gene Identification

When applying stringent thresholds (MAPQ ≥ 30 for alignment-based methods and a confidence score of 0.8 for Kraken2), we observed that bowtie2 (very_sensitive_local mode) was the optimal aligner in 9 out of 35 cases, followed by bowtie2(sensitive and very sensitive modes). These results highlight the robustness of Bowtie2 (very_sensitive_local mode) under stringent quality criteria, suggesting it provides the most reliable alignments for cfDNA-based AMR gene detection in our benchmarking framework.

### (10) Result of AMR gene Alignment pre/post filtering

Before human read removal, 35 samples had at least one AMR gene present, while after human read decontamination step, 34 out of 102 samples had at least one AMR gene present. Unlike bacterial species, the presence of anti-microbial genes was constant in 24 samples before and after the human read removal step. Only ten samples after the human read removal step saw a change in AMR gene spectrum compared to when no human reads were removed.

Before human read removal, the most frequent AMR genes were APH(3)-Ib in *Pseudomonas aeruginosa* (8 samples), APH(6)-Id in *Pseudomonas aeruginosa* (6 samples), and tet(M) in *Erysipelothrix rhusiopathiae* (4 samples). After filtering, APH(3)-Ib increased to 13 samples, while APH(6)-Id (6 samples) and tet(M) (3 samples) remained unchanged. Detailed pre- and post-filtering frequencies are provided in the Supplementary Data.

## Discussions

Plasma cell-free DNA (cfDNA) profiles in sepsis are dominated by human reads, making accurate filtering a critical prerequisite for metagenomic analysis. The filtering process must be both precise and stringent to prevent human reads from inflating bacterial and antimicrobial resistance (AMR) abundances by misaligning to microbial or AMR gene reference genomes. However, excessive stringency also risks discarding genuine bacterial and AMR gene reads. Thus, a careful balance between human read removal and microbial read retention must be evaluated for each aligner before it is applied to cfDNA studies.

To identify the optimal aligner for human read removal, we evaluated the retention capability of six aligners using in silico cfDNA profiles generated from pure bacterial FASTA files. We then assessed the stringency and specificity of each aligner in patient cfDNA samples. These analyses demonstrated that BWA-MEM2 performed best in both stringency and specificity, despite having the lowest retention of microbial reads. Our choice is guided by the findings of Gihawi et al^7^, who highlighted the risks of human reads being misaligned to bacterial reference genomes, which can inflate microbial profiles and generate false positives. Therefore, an ideal aligner should minimize false positives, even at the expense of losing a few genuine microbial reads. Using an aligner with high retention but poor human-read removal could artificially broaden the microbial spectrum, complicating the distinction of true signals from false positives. Considering these trade-offs, BWA-MEM2 emerges as the preferred aligner for human read removal in cfDNA metagenomic analyses.

It is important to note that in cfDNA samples generated with highly careful library preparation, where the proportion of human reads is already minimal, retention may be a more critical parameter than stringency or specificity, and thus the optimal aligner could differ. However, in our cohort, where >99% of reads were human, retention was of lower priority, and stringency and specificity became the decisive factors.

To illustrate the BWA-MEM2 advantage over its close competitor Bowtie2 (very-sensitive-local) for cleaning human reads, we generated two in silico cfDNA datasets from the human CHM13-T2T reference: one with an average of 0.03427 million residual reads (average human reads found in Table 2 after BWA-MEM2 cleaning) and another with 0.05961 million residual reads (average human reads found in Table 2 after Bowtie2 very-sensitive-local cleaning). When both were aligned to the Combined D1 Filtered Bacterial Sepsis Database using BWA-MEM2 (choice of post-cleaning bacterial taxa aligner), the Bowtie2-cleaned set produced 69% more spurious bacterial mappings, reflecting inflated profiles due to misclassified human reads. This underscores the risk of false positives with Bowtie2 (very-sensitive-local) and highlights BWA-MEM2 as the more reliable tool for human read removal in cfDNA analysis.

For antimicrobial resistance (AMR) gene detection, Bowtie2 in very-sensitive-local mode ranked first, yielding the highest ratio of true to false hits, followed by other Bowtie2 modes. In our 35 in silico cfDNA profiles, no single aligner consistently outperformed others in more than half of the cases. However, the top three performers were all Bowtie2 variants, with the very-sensitive-local mode emerging as the most reliable for AMR gene detection. Because very-sensitive-local mode uses more permissive alignment parameters (e.g., shorter seed length and local alignment with soft clipping), it is likely to capture more true AMR gene hits compared to less-sensitive modes.

Our criteria for reporting a true bacterial hit required reads to align to complete genomes, excluding plasmids. Although certain host-associated plasmids can, in principle, provide evidence of species presence, we did not include them in this analysis. Given the vast number of plasmids and the complexity of distinguishing those with strong host specificity from broadly mobile elements, reliably incorporating plasmid-based signals into species identification remains challenging.

The accuracy of aligners depends heavily on the reference genome database used. To minimize read misallocation and reduce the “read wastage” phenomenon reported with large public databases, we constructed a curated reference set containing only species relevant to sepsis.

## Conclusion

BWA-MEM2 emerged as the most effective aligner for human read removal, balancing stringency and specificity, though it occasionally removed genuine microbial reads. It also outperformed other aligners in bacterial taxonomic classification by aligning reads more accurately to complete bacterial genomes. For antimicrobial resistance gene detection from human-read–filtered cfDNA paired-end profiles, Bowtie2 in very-sensitive-local mode proved to be the preferred choice. We hope that these bioinformatic alignment guidelines will enable more accurate identification of plasma cfDNA microbiome and AMR genes profiles, thereby assisting clinicians in sepsis diagnosis and in making informed antibiotic treatment decisions.

## Study Limitations and Future Directions

Benchmarking relied on in-silico simulations rather than synthetic cfDNA controls, which may not fully capture the complexity of in vivo cfDNA, where microbial fragments vary in size, abundance, and degradation patterns. Consequently, aligners may behave differently on real patient samples, affecting human read removal, bacterial retention, and AMR gene detection.

All samples were derived from an acute leukemia (AL) cohort; therefore, our observations and inferences regarding human read removal may not be valid for other cancer subtypes due to the nature of cell-free DNA fragments. Future testing across pan-cancer and non-cancer cfDNA plasma samples could help evaluate the broader applicability of our findings.

Our findings are based on paired cfdna data and not on single-end cfdna, which would have different optimal aligner outcomes in both filtering and bacterial/AMR gene alignment.

Our metagenomic analyses were highly sensitive but not specific to individual species or AMR genes, making it difficult to identify dominant organisms as is typically done in standard blood cultures. Due to limited sequencing depth, we were unable to generate Metagenome-Assembled Genomes (MAGs) from these samples. In this study, we benchmarked six widely used aligners, most of which are read-based, with only one representing a k-mer–based approach. Other alignment strategies—including pseudo-aligners, marker-gene classifiers, assembly-based approaches, and graph-based aligners—were not included. Including such methods in future benchmarking could provide a more comprehensive evaluation of alignment performance in plasma cfDNA metagenomics.

We used the T2T-CHM13 genome for host read filtration, as it resolves missing Y-chromosomal and centromeric regions, reducing false microbial assignments. While the HPRC pangenome^35^ offers broader population and structural diversity and could further improve human read removal, T2T-CHM13 was sufficient for cfDNA analyses in this study.

Our analysis was limited only to bacterial and AMR Gene identification, and our optimal aligners cannot be generalized for other infections, such as fungal and viral.

## Supporting information

Supplemental Excels

## Data Availability

The raw FASTQ sequencing data generated and analyzed in this study are available from the corresponding author or principal investigator upon reasonable request.

## Ethics Statement

Institutional Ethics Committee III (TMC/IEC III) has given approval for this study (Project Number: 900820) dated 30th June 2021.

## Supplementary Sheet link

Supplementary Excel

## Authors Contribution

### Study Design and Hypothesis

Nikhil Patkar, Anant Gokarn

### Data Analysis and Interpretation

Arpit Mathur, Vishram Terse, Nikhil Patkar, Anant Gokarn, Sujata Lall, Elwin Paulose

### Wet Lab Preparation

Karishma Anam, Prasanna Bhanshe, Swapnali Joshi, Shruti Chaudhary, Manisha More, Pratiksha Salunke

### Sepsis Data Curation

Sujata Lall, Vivek Bhatt, Arpit Mathur

### Data Management

Arpit Mathur, Vaibhav Gawde, Moksha Dhoka

### Clinical Support (Patient Recruitment and Counseling)

Prashant Tembhare, Sweta Rajpal, Gaurav Chatterjee, Papagudi Ganesan Subramanian, Sumeet Gujral, Sachin Punatar, Summet Mirgh, Akanksha Chichra, Gaurav Bain, Navin Khattry, Anant Gokarn, Manju Sengar, Alok Shetty, Hasmukh Jain, Lingaraj Nayak, Bhausaheb Bagal.

